# Intrauterine Adhesions and Prior use of a Progestin-releasing Intrauterine Device

**DOI:** 10.64898/2026.06.24.26356491

**Authors:** Kaia Schwartz, Alyssa Zhou, Jennifer Aranda, Nikhita Mahendru, Caroline Hodge, David Huang, Elena HogenEsch, Heather Huddleston

## Abstract

**Objective:** To determine whether prior progestin-releasing intrauterine device (progestin-IUD) use is associated with intrauterine adhesions (IUA).

**Design:** Retrospective case-control study

**Subjects:** Women evaluated at a single academic reproductive endocrinology practice, including sequential patients with: (a) hysteroscopically confirmed intrauterine adhesions (IUA cases; n=371), (b) hysteroscopically confirmed endometrial polyps (polyp control group; n=348), or (c) a new patient visit for male-factor or unexplained infertility (infertile comparison group; n= 757).

**Exposure:** Self-reported prior contraceptive use, ascertained from structured intake forms completed prior to the initial visit; verified by chart review

**Main Outcome Measures:** Adjusted odds ratios (aOR) for IUA associated with progestin-IUD use, from logistic regression adjusting for age, calendar year, race/ethnicity, prior dilation and curettage/manual uterine aspiration, operative hysteroscopy, myomectomy, and prior live birth.

**Results:** Progestin-IUD use was more frequent among IUA cases (29.9%) than polyp (3.4%) or fertility (11.1%) comparison groups. Compared to infertile comparators, any prior progestin-IUD use was independently associated with IUA (aOR 3.12; 95% CI 2.01–4.85). There was a duration-response pattern: use ≤5 years of total use was modestly associated with IUA case status (aOR 1.99; 1.09–3.64), whereas use >5 years of total use conferred more than a sevenfold increase (aOR 7.26; 3.27–16.11). The association persisted among surgically naïve women (aOR 3.98; 2.44–6.48) and was concentrated in those who were nulliparous, where use beyond five years conferred an approximately twelve-fold increase in odds (aOR 12.74; 5.25–30.92). Progestin-IUD use was inversely associated with polyp controls relative to IUA and infertile controls.

**Conclusion:** Prior progestin-IUD use, particularly beyond five years and among nulliparous women, was independently associated with IUA across two distinct comparison groups, warranting future prospective studies to evaluate reproductive outcomes after long-term use.

## Introduction

Intrauterine adhesions (IUA) are characterized by the formation of fibrous tissue within the uterine cavity following endometrial trauma. IUA, often referred to as Asherman’s syndrome, affects an estimated 1.5% of reproductive-age women and is associated with significant reproductive morbidity including infertility, recurrent pregnancy loss, and menstrual irregularities(1). Hysteroscopy remains the gold standard for both diagnosis and treatment, though patients often continue to experience adverse reproductive outcomes despite intervention(1). While pregnancy-related uterine instrumentation, particularly dilation and curettage, represents the most well-established risk factor, causes unrelated to pregnancy, including hysteroscopic procedures and endometrial inflammation, have also been implicated. Despite its clinical relevance, the full spectrum of risk factors contributing to adhesion formation remains incompletely understood, and opportunities for primary prevention are limited.

Intrauterine devices (IUD’s) have a high degree of contraceptive efficacy and have become a widely used choice over the past two decades(2). A 2023 report using data from the National Survey of Family Growth estimated that 15% of all sexually active women ages 15-49 had ever used a hormonal IUD. Return to fertility guidance following progestin-IUD use was initially founded on fertility rates for women in contraceptive efficacy studies who discontinued early to conceive. In three studies where IUD was less than 5 years, normal conception rates were reported for a mix of nulliparous and parous women (total n for all studies = 385)(3–5). However the equivalent study reporting reassuring fertility rates for women who used the device for more than five years included only 31 women who were seeking pregnancy(6). Subsequent large prospective studies have also indicated reassuring fecundability and miscarriage rates for hormonal IUD, although there is a lack of studies tracking long-term use specifically and none have examined post-miscarriage sequelae(7, 8).

Against the backdrop of reassuring fecundability and miscarriage data, emerging clinical observations have suggested that progestin-intrauterine devices might represent an independent risk factor for IUA (9, 10). Unfortunately, to date, these observations are lacking a strong evidence base. Given the widespread use of progestin-IUDs among reproductive-age women and the excellent contraceptive efficacy, it is critical that this question be examined in a robust manner. Accordingly, in the present study we sought to evaluate potential risk factors for IUA formation using a retrospective case-control design, with a specific focus on prior contraceptive history including progestin-IUD use.

## Methods

### Study Design and Population

We conducted a retrospective case control study comparing contraceptive history among three groups: women diagnosed at hysteroscopy with intrauterine adhesions (IUA) (cases), women diagnosed at hysteroscopy with endometrial polyps (polyp control group), and women diagnosed with male or unexplained infertility (infertile control group), all seen at the Center for Reproductive Health (CRH), a single academic reproductive endocrinology practice at the University of California, San Francisco (UCSF) between 2015 and 2024. The study was approved by the UCSF Institutional Review Board.

The surgical groups were identified from a list generated by electronic health record (EHR) of all hysteroscopic procedures performed at CRH between 2015-2024. Manual chart review was performed to confirm diagnoses, pathology and other variables related to the procedure. For patients with multiple procedures, only the first procedure was considered (index procedure). The **IUA group** comprised 371 sequential patients with a documented diagnosis of intrauterine adhesions who underwent hysteroscopic lysis as the index procedure. The **polyp group** comprised 348 women undergoing hysteroscopic polypectomy as the index procedure. Due to the large volume of this procedure, only sequential procedures performed between 2015 and 2020 were reviewed. The **infertile control group** was generated from an EHR-generated list of sequential patients with a diagnosis of either unexplained or male factor infertility (and therefore a non-uterine fertility diagnosis), based on ICD-10 codes, seen at CRH. We included the first 100 patients per year for each year of the study period (2017-2024). Manual chart review was performed to confirm fertility diagnoses. Patients were excluded if their visit reason was fertility preservation or desire for donor sperm insemination (same-sex and single patients). Patients were also excluded if they had any history of hysteroscopic intervention for uterine adhesions.

### Exposure Ascertainment

Contraceptive history was ascertained from manual review of data collected from structured intake forms administered when patients first presented for care. Critically, the form asks new patients to report all contraceptives used in the past and dates of use. Additional manual chart review was performed to verify intake data. The primary exposure was prior progestin-releasing IUD use, defined as self-reported use of any progestin IUD (Mirena, Kyleena, Liletta, or Skyla) prior to the index procedure. Duration of progestin IUD use was recorded in months and categorized as: IUD use 1 to ≤60 months (≤5 years); IUD use >60 months (>5 years); or IUD use with missing or zero duration recorded (duration unknown). Total months of use were considered. Additional contraceptive methods were also ascertained as a secondary exposure.

### Covariate Ascertainment

Prior pregnancy history was collected from new patient intake forms or manual chart review and included information on gravidity, live births, spontaneous abortions (SAB), therapeutic abortions (TAB, recorded separately as medical and surgical), biochemical pregnancies, ectopic pregnancies, and intrauterine fetal demise. Additional demographic and surgical history details were similarly obtained from intake forms.

### Statistical Analysis

Baseline characteristics were compared using the Kruskal–Wallis test for continuous variables and the chi-square test for categorical variables. The association between progestin IUD use and group membership was examined using logistic regression across three pairwise comparisons: IUA versus Infertility Control, Polyp Control versus Infertility Control, and IUA versus Polyp Control. Both unadjusted odds ratios (OR) and adjusted odds ratios (aOR) with 95% confidence intervals (CI) are reported.

Exposures included: progestin-IUD (any); progestin-IUD only (no reported use of OCP); reported use of both OCP and progestin-IUD in the past; IUD ≤5 years; IUD >5 years; IUD (duration unknown; users with missing or zero duration recorded); OCP and any other method, and OCP only. The fully adjusted model included factors identified a priori as potential confounders and/or those that differed across groups at baseline, including: age (continuous, mean-centered), calendar year of index surgery (IUA or polyp controls) or new patient visit (infertile control) (centered at 2020), race/ethnicity (White [reference] / Asian / Other), prior D&C/manual uterine aspiration (MUA) (binary), prior operative hysteroscopy (binary), prior myomectomy (binary), and prior live birth (binary).

Two pre-specified sensitivity analyses were conducted for the primary IUA vs Infertile control comparisons. First, models were stratified by prior uterine surgery (surgically naïve vs any prior procedure, inclusive of hysteroscopy, D&C/Manual uterine aspiration (MUA), or myomectomy); surgery indicators were omitted from the covariate set in the surgically naïve stratum. Second, models were stratified by prior live birth (live birth=0 vs live birth ≥1); prior live birth was omitted from covariates within strata.

All analyses were performed in Stata using logistic regression with unpenalized maximum likelihood estimation. Statistical significance was defined as two-sided p<0.05.

## RESULTS

### Study Population

A total of 1,476 women were included: 371 with IUA, 348 with endometrial polyps, and 757 with a non-uterine infertility diagnosis (Table 1). IUA patients were older than infertile controls (mean age 38.4 vs 36.7 years; p<0.001) but similar in age to polyp patients (38.4 years). Gravidity was substantially higher in the IUA group than in either comparison group (mean [SD] 2.2 [2.2] vs 0.7 [1.2] in polyp and 0.7 [1.2] in infertile controls; p<0.001), whereas the polyp and infertile groups did not differ from one another. Prior dilation and curettage or manual uterine aspiration (D&C/MUA) was reported by 54.7% of IUA patients, compared with 10.1% of polyp patients and 2.8% of infertile controls (p<0.001). The IUA group also had higher rates of any prior spontaneous abortion (50.1% vs 15.8% and 15.2%; p<0.001) and any prior therapeutic abortion (27.0% vs 11.2% and 11.9%; p<0.001), with similar rates in the two comparison groups. Asian women comprised 44.5% of the polyp group, compared with 27.2% of the IUA group and 31.3% of controls (p<0.001).

**Table 1.** Baseline characteristics of the study cohort, with overall and pairwise comparisons.

| Characteristic | IUA (n=371) | Polyp Control (n=348) | Infertile Control (n=757) | Overall p | IUA vs Infertile Control | Polyp Control vs Infertile Control | IUA vs Polyp |
| --- | --- | --- | --- | --- | --- | --- | --- |
| **Demographics** |  |  |  |  |  |  |  |
| Age, years, mean (SD) | 38.4 (4.2) | 38.4 (4.6) | 36.7 (4.1) | <0.001 | <0.001 | <0.001 | 0.855 |
| BMI, kg/m<sup>2</sup>, mean (SD) | 24.7 (4.7) | 24.4 (4.9) | 25.3 (5.8) | 0.105 | 0.756 | 0.053 | 0.070 |
| Year of procedure, median (range) | 2021 (2016–2024) | 2019 (2015–2022) | 2021 (2017–2024) | <0.001 | 0.004 | <0.001 | <0.001 |
| Race/ethnicity, n (%)† |  |  |  | <0.001 | 0.002 | <0.001 | <0.001 |
| White | 168 (45.3) | 117 (33.6) | 323 (42.7) |  |  |  |  |
| Asian | 101 (27.2) | 155 (44.5) | 237 (31.3) |  |  |  |  |
| Hispanic | 40 (10.8) | 25 (7.2) | 44 (5.8) |  |  |  |  |
| Black | 8 (2.2) | 4 (1.1) | 16 (2.1) |  |  |  |  |
| Mixed | 13 (3.5) | 3 (0.9) | 63 (8.3) |  |  |  |  |
| Unknown/not reported | 41 (11.1) | 44 (12.6) | 74 (9.8) |  |  |  |  |
| **IUA Severity (Intrauterine adhesions group only)** |  |  |  |  |  |  |  |
| Mild (Grade 1), n (%) | 230 (62.0) | — | — |  |  |  |  |
| Moderate (Grade 2), n (%) | 112 (30.2) | — | — |  |  |  |  |
| Severe (Grade 3), n (%) | 29 (7.8) | — | — |  |  |  |  |
| **Prior Obstetric History** |  |  |  |  |  |  |  |
| Prior pregnancies (G), mean (SD)‡ | 2.2 (2.2) | 0.7 (1.2) | 0.7 (1.2) | <0.001 | <0.001§ | 0.360§ | <0.001§ |
| G=0, n (%) | 56 (15.1) | 215 (61.8) | 431 (56.9) |  |  |  |  |
| G=1, n (%) | 101 (27.2) | 61 (17.5) | 155 (20.5) |  |  |  |  |
| G≥2, n (%) | 208 (56.1) | 67 (19.3) | 133 (17.6) |  |  |  |  |
| Prior live births (L), mean (SD) | 0.6 (0.7) | 0.2 (0.6) | 0.2 (0.5) | <0.001 | <0.001§ | 0.521§ | <0.001§ |
| L=0, n (%) | 189 (50.9) | 278 (79.9) | 566 (74.8) |  |  |  |  |
| L=1, n (%) | 152 (41.0) | 49 (14.1) | 123 (16.2) |  |  |  |  |
| L≥2, n (%) | 24 (6.5) | 12 (3.4) | 26 (3.4) |  |  |  |  |
| Any prior SAB, n (%) | 186 (50.1) | 55 (15.8) | 115 (15.2) | <0.001 | <0.001 | 0.788 | <0.001 |
| Any prior TAB, n (%) | 100 (27.0) | 39 (11.2) | 90 (11.9) | <0.001 | <0.001 | 0.840 | <0.001 |
| **Prior Surgical History** |  |  |  |  |  |  |  |
| D&C or MUA, n (%) | 203 (54.7) | 35 (10.1) | 21 (2.8) | <0.001 | <0.001 | <0.001 | <0.001 |
| Prior operative hysteroscopy, n (%) | 50 (13.5) | 35 (10.1) | 27 (3.6) | <0.001 | <0.001 | <0.001 | 0.167 |
| Myomectomy, n (%) | 22 (5.9) | 8 (2.3) | 9 (1.2) | <0.001 | <0.001 | 0.190 | 0.016 |
| No prior surgery, n (%) | 129 (34.8) | 275 (79.0) | 701 (92.6) | <0.001 | <0.001 | <0.001 | <0.001 |
| **Contraceptive History** |  |  |  |  |  |  |  |
| Combined oral contraceptives, n (%) | 244 (65.8) | 221 (63.5) | 431 (56.9) | 0.008 | 0.004 | 0.041 | 0.533 |
| Progestin-IUD (any), n (%) | 111 (29.9) | 12 (3.4) | 84 (11.1) | <0.001 | <0.001 | <0.001 | <0.001 |
| Duration, mean (SD), months** | 61.3 (40.4) | 43.0 (48.0) | 45.8 (38.8) | 0.029 | 0.020 | 0.541 | 0.085 |
| Median (IQR), months | 50 (24–96) | 36 (5–54) | 36 (12–60) |  |  |  |  |
| Total use ≤5 years, n (%) | 49 (13.2) | 9 (2.6) | 47 (6.2) | <0.001 | <0.001 | 0.011 | <0.001 |
| Total use >5 years, n (%) | 31 (8.4) | 2 (0.6) | 15 (2.0) | <0.001 | <0.001 | 0.112 | <0.001 |
| Duration unknown, n (%) | 31 (8.4) | 1 (0.3) | 22 (2.9) | <0.001 | <0.001 | 0.003 | <0.001 |
| Copper IUD, n (%) | 18 (4.9) | 7 (2.0) | 24 (3.2) | 0.108 | 0.181 | 0.331 | 0.042 |
| Depot Provera (DMPA), n (%) | 16 (4.3) | 6 (1.7) | 26 (3.4) | 0.136 | 0.504 | 0.126 | 0.052 |
| NuvaRing or patch, n (%) | 23 (6.2) | 14 (4.0) | 29 (3.8) | 0.175 | 0.095 | 0.868 | 0.237 |
| Progestin-only pill, n (%) | 0 (0.0) | 2 (0.6) | 6 (0.8) | 0.233 | 0.186 | 1.000 | 0.234 |
| Nexplanon implant, n (%) | 5 (1.3) | 0 (0.0) | 9 (1.2) | 0.109 | 0.782 | 0.064 | 0.062 |

**p-values:** Overall p compares all three groups (Kruskal–Wallis for continuous variables; chi-square for categorical variables). Pairwise p-values are post-hoc two-group comparisons: Fisher’s exact test for binary variables, chi-square for multi-category variables (race/ethnicity), and Mann–Whitney U for continuous variables.
† Race/ethnicity: IUA and Polyp groups used a standardised OMB dropdown at intake; Control group race was derived from the EHR free-text primary ancestry field. Pairwise comparisons are chi-square tests across the displayed categories.
‡ Prior pregnancies (G) include live births, spontaneous abortions, therapeutic abortions, biochemical pregnancies, ectopic pregnancies, and stillbirths. Missing: IUA n=6, Polyp n=5, Control n=38. Parity unknown in 57 patients (6 IUA, 9 polyp, 42 control), coded as nulliparous in adjusted models.
¶ Surgery indicators are not mutually exclusive. D&C includes dilation and curettage or manual uterine aspiration for any indication. Operative hysteroscopy excludes the index procedure. No prior surgery indicates absence of all listed procedures.
\*\* Duration restricted to IUD users with recorded duration >0 months (IUA n=80/111, Polyp Control n=11/12, Infertile Control n=62/84). Duration unknown: IUD users with missing or zero duration recorded (shown in *italic*).

### Contraceptive use

Progestin-IUD use was reported by a significantly higher proportion of IUA patients (29.9%) than of polyp (3.4%) or infertile (11.1%) controls; use was also lower in the polyp group than among infertile controls (overall p<0.001)(Table 1). Among users with a recorded duration, median use was longer in the IUA group (50 months; IQR 24–96) than in the polyp (36 months; IQR 5–54) or infertile (36 months; IQR 12–60) groups (p=0.029); duration was missing or recorded as zero in 31 IUA, 1 polyp, and 22 infertile patients. Combined oral contraceptive use was reported by 65.8% of IUA patients, 63.5% of polyp controls, and 56.9% of infertile controls (p=0.008). Prior use of the copper IUD, depot medroxyprogesterone acetate, and the contraceptive ring or patch did not differ across groups, and use of these methods was uniformly low.

Although the endometrial polyp group was initially planned as a surgical comparator, progestin-IUD use was found to be markedly lower among polyp patients (3.4%) compared to infertile controls (11.1%) and was also significantly lower than population estimates for women ages 15-44 (∼15%)(11). This finding could be consistent with progestin-mediated endometrial suppression reducing polyp formation (adjusted OR for IUD use, polyp vs control, 0.39; 95% CI 0.20–0.76). Because the exposure itself likely influences membership in the polyp group, we determined that this cohort did not constitute a neutral reference for the exposure. As such, the infertile control comparison was designated as primary, with the polyp controls reported as secondary

### Main Analysis

Table 2 and Figure 1 present adjusted and unadjusted odds ratios for prior progestin-IUD use, combined progestin-IUD and combined oral contraceptive pill (OCP) use, any OCP use, and OCP use alone. In the fully adjusted model, prior progestin-IUD use was associated with significantly increased odds of IUA compared with infertile controls (aOR 3.12, 95% CI 2.01–4.85; p<0.001) (Table 2, Figure 1). A duration-response gradient was observed: use of ≤5 years was modestly associated with IUA (aOR 1.99, 95% CI 1.09–3.64; p=0.025), whereas use of >5 years was associated with a more than seven-fold increase in odds (aOR 7.26, 95% CI 3.27–16.11; p<0.001). Users with unknown duration showed an intermediate, significant association (aOR 3.30, 95% CI 1.55–7.02; p=0.002). The IUA group was likewise associated with markedly increased odds of progestin-IUD use versus polyp controls (aOR 7.08, 95% CI 3.43–14.64; p<0.001). The polyp group and a lower odds of progestin-IUD use versus infertile controls (aOR 0.39, 95% CI 0.20–0.76; p=0.006).

**Figure 1:**
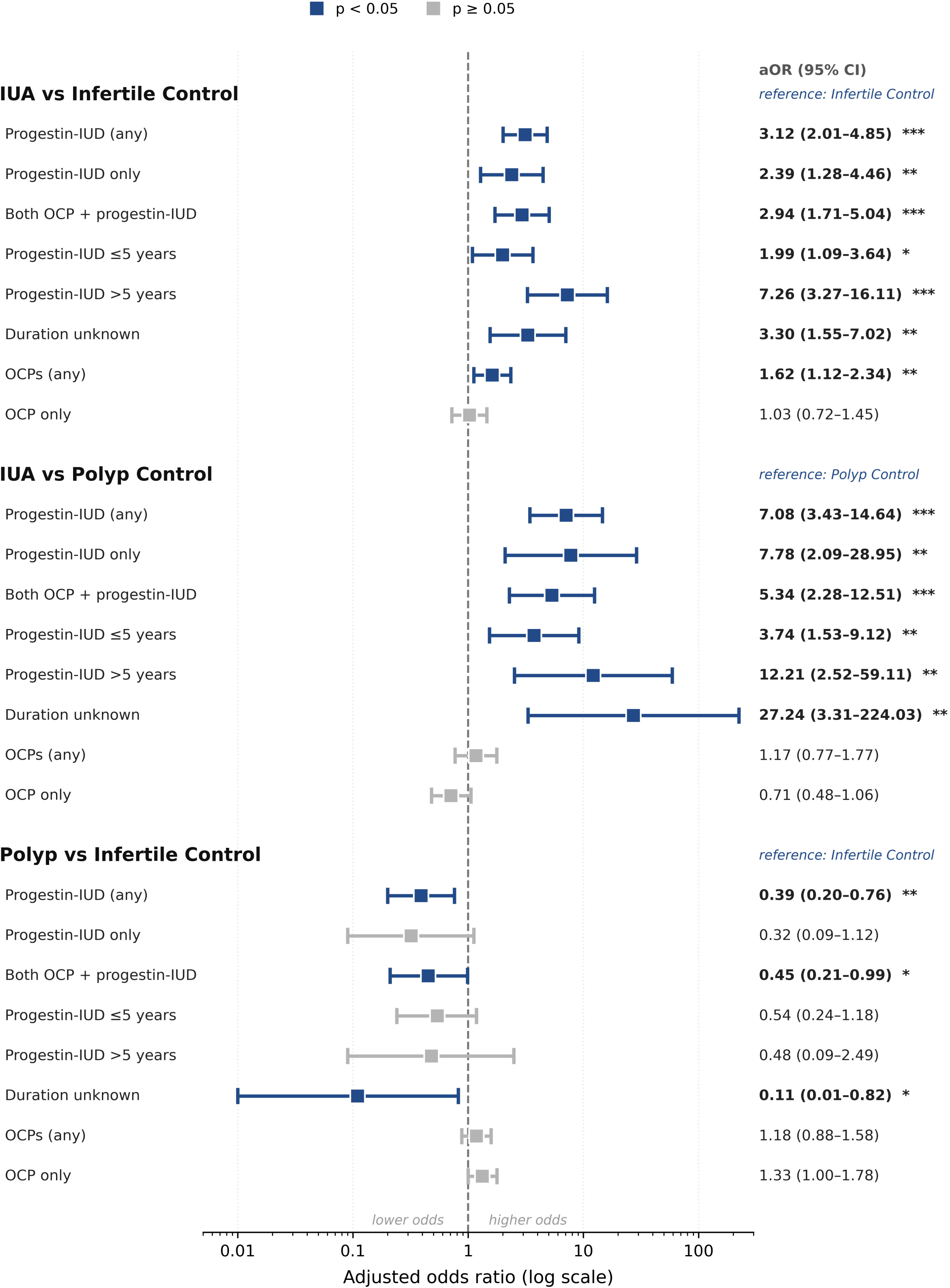
Association of prior contraceptive use with intrauterine adhesions.

**Table 2.** Unadjusted and adjusted odds ratios for prior contraceptive use across the three pairwise comparisons.

|  | IUA vs Infertile Control |  | Polyp Control vs Infertile Control |  | IUA vs Polyp Control |  |
| --- | --- | --- | --- | --- | --- | --- |
| Exposure | uOR (95% CI) | aOR (95% CI) | uOR (95% CI) | aOR (95% CI) | uOR (95% CI) | aOR (95% CI) |
| ***Progestin- IUD*** |  |  |  |  |  |  |
| IUD (any) | 3.42 (2.49–4.70)*** | **3.12 (2.01–4.85)\*\*\*** | 0.29 (0.15–0.53)*** | **0.39 (0.20–0.76)\*\*** | 11.95 (6.45–22.16)*** | **7.08 (3.43–14.64)\*\*\*** |
| IUD only | 3.01 (1.90–4.78)*** | **2.39 (1.28–4.46)\*\*** | 0.18 (0.06–0.61)** | 0.32 (0.09–1.12)† | 16.28 (5.01–52.85)*** | **7.78 (2.09–28.95)\*\*** |
| Both OCP + IUD | 3.00 (2.03–4.45)*** | **2.94 (1.71–5.04)\*\*\*** | 0.38 (0.18–0.77)** | **0.45 (0.21–0.99)\*** | 8.00 (3.92–16.34)*** | **5.34 (2.28–12.51)\*\*\*** |
| IUD ≤5 years | 2.70 (1.76–4.13)*** | **1.99 (1.09–3.64)\*** | 0.38 (0.19–0.79)** | 0.54 (0.24–1.18) | 7.04 (3.39–14.59)*** | **3.74 (1.53–9.12)\*\*** |
| IUD >5 years | 5.35 (2.84–10.07)*** | **7.26 (3.27–16.11)\*\*\*** | 0.27 (0.06–1.17)† | 0.48 (0.09–2.49) | 20.03 (4.75–84.46)*** | **12.21 (2.52–59.11)\*\*** |
| IUD (duration unknown) | 3.65 (2.07–6.42)*** | **3.30 (1.55–7.02)\*\*** | 0.09 (0.01–0.68)* | **0.11 (0.01–0.82)\*** | 40.06 (5.43–295.39)*** | **27.24 (3.31–224.03)\*\*** |
| ***Combined oral contraceptives*** |  |  |  |  |  |  |
| OCPs (any) | 1.45 (1.12–1.88)** | **1.62 (1.12–2.34)\*\*** | 1.32 (1.01–1.71)* | 1.18 (0.88–1.58) | 1.10 (0.81–1.50) | 1.17 (0.77–1.77) |
| OCP only | 0.92 (0.72–1.18) | 1.03 (0.72–1.45) | 1.54 (1.19–1.99)** | 1.33 (1.00–1.78)† | 0.60 (0.44–0.80)*** | 0.71 (0.48–1.06)† |

Each cell is a separate logistic regression. uOR, unadjusted odds ratio; aOR, adjusted odds ratio (age, calendar year, race/ethnicity [White/Asian/Other], prior D&C/MUA, operative hysteroscopy, myomectomy, prior live birth). IUD duration categories are modeled versus never-use of a progestin-IUD. “IUD only,” “Both OCP + IUD,” and “OCP only” are each modeled as that exposure group versus all other women. Oral contraceptive (OCP)
†p<0.10; \*p<0.05; \*\*p<0.01; \*\*\*p<0.001. Cohort: IUA n=371, Polyp n=348, infertile control n=757. Complete-case analytic n=1,128 (IUA vs control), 718 (IUA vs polyp), 1,104 (polyp vs control).

Any prior OCP use was also associated with increased odds of IUA versus the infertility comparator (aOR 1.62, 95% CI 1.12–2.34; p=0.010). However, this association was fully attenuated among women who had used OCPs but never a progestin-IUD (aOR 1.03, 95% CI 0.72–1.45; p=0.886), whereas women who had used both an OCP and a progestin-IUD had elevated odds comparable to progestin-IUD users overall (aOR 2.94, 95% CI 1.71–5.04; p<0.001). The OCP association therefore appeared to be confounded by progestin-IUD use rather than reflective of an independent effect of OCPs.

### Other Contraceptives

No other contraceptive methods were significantly associated with IUA. Notably, the copper IUD—which exerts no hormonal effect on the endometrium—showed no association (aOR 1.17, 95% CI 0.57–2.40), whereas estimates for depot medroxyprogesterone (aOR 1.72, 95% CI 0.74–4.02) and the contraceptive ring or patch (aOR 1.79, 95% CI 0.87–3.72) were non-significant. Limited utilization of the etonogestrel implant and progestin only pill prevented determination of reliable estimates.

### Covariate Associations

Prior D&C/MUA was the strongest independent predictor of IUA (aOR 40.59, 95% CI 24.46–67.36; p<0.001), followed by prior myomectomy (aOR 11.31, 9-% CI 4.66–27.45; p<0.001) and operative hysteroscopy (aOR 4.03, 95% CI 2.18–7.43; p<0.001). Prior live birth was positively associated with IUA (aOR 3.06, 95% CI 2.12–4.42; p<0.001), likely reflecting the higher burden of uterine procedures in multiparous women rather than a causal relationship. Race/ethnicity was not independently associated with IUA.

### Sensitivity Analyses

Results of both sensitivity analyses are summarized in *Table 3* and *Figure 2*. Among surgically naïve patients (n=129 IUA, n=701 controls), the progestin-IUD-IUA association was robust overall: aOR 3.98 (95% CI 2.44–6.48; p<0.001), with a higher magnitude in those with prior use >5 years: aOR 10.18 (95% CI 4.38–23.69; p<0.001). Among patients with unknown IUD duration, the aOR was 4.00 (95% CI 1.77–9.03; p<0.001) in this stratum. Estimates in the prior surgery stratum were non-significant across all IUD categories, but this stratum was severely limited by a small control group (n=56 controls with any prior surgery), and results should be interpreted with caution.

**Figure 2:**
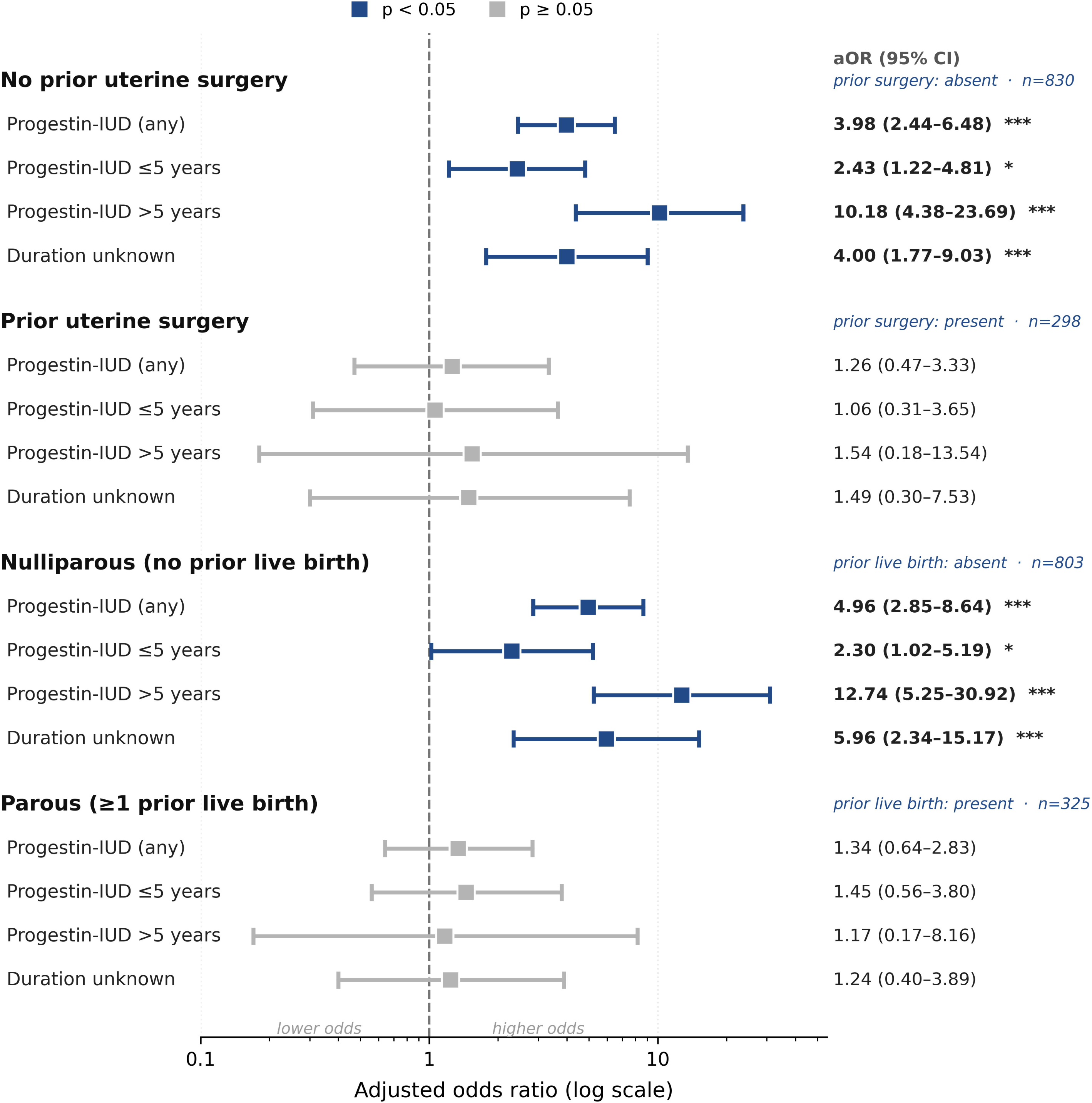
Progestin-lUD and intrauterine adhesions, stratified analyses (IUA vs Infertile Control)

**Table 3.**
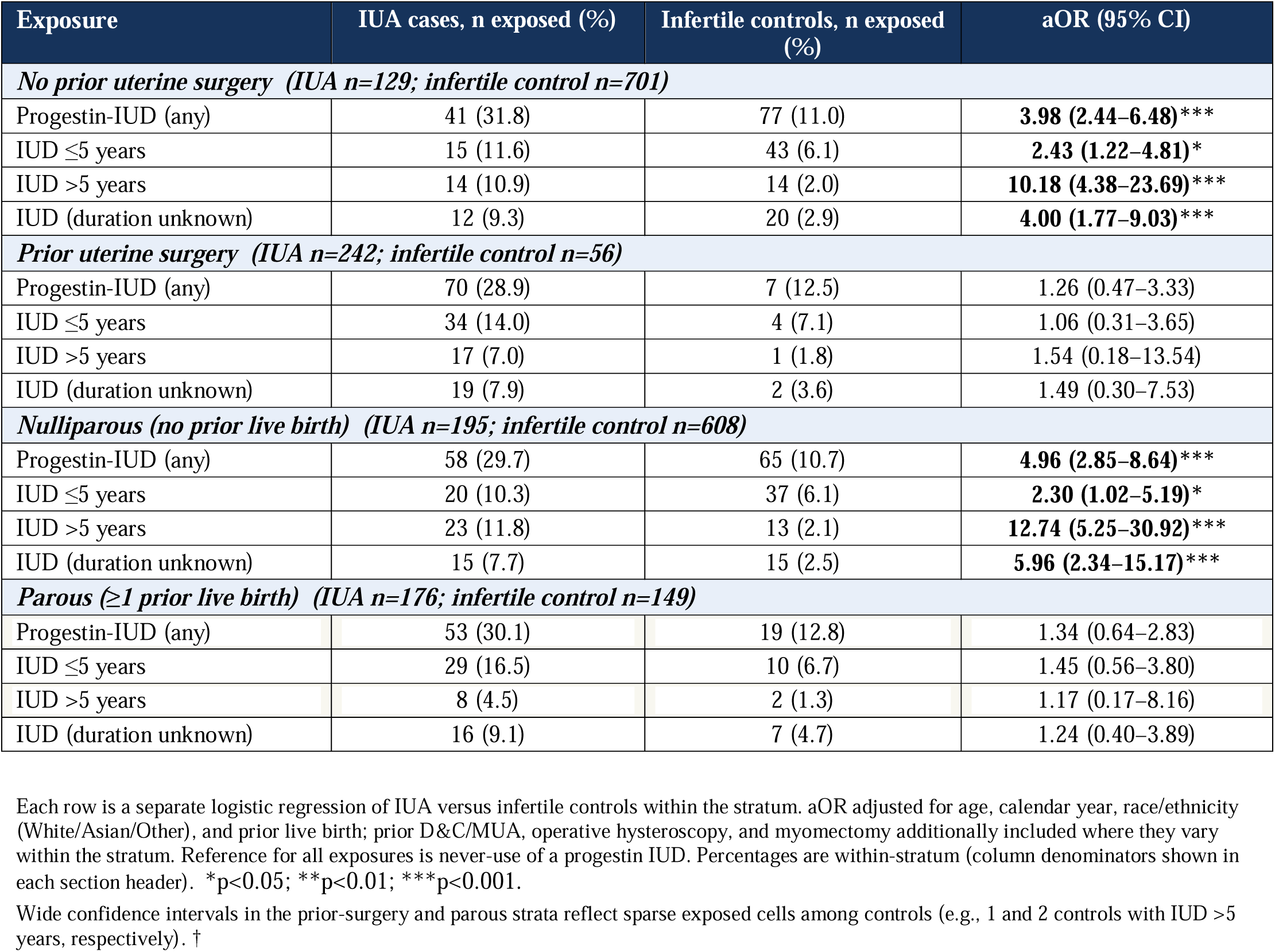
Progestin-IUD use and intrauterine adhesions (IUA vs infertile controls), stratified by prior uterine surgery and by prior live birth, with exposure counts.

Stratified by parity, the IUD-IUA association was concentrated in nulliparous women (live birth=0; n=195 IUA, n=608 controls): aOR 4.96 (95% CI 2.85–8.64; p<0.001) for any IUD, and aOR 12.74 (95% CI 5.25–30.92; p<0.001) for IUD >5 years. Women with unknown IUD duration also showed a significant association in the nulliparous stratum (aOR 5.96, 95% CI 2.34–15.17; p<0.001). Among parous women (live birth≥1), all progestin-IUD associations were attenuated and non-significant.

## DISCUSSION

In this retrospective case–control study, prior use of a progestin-containing intrauterine device (IUD) was strongly associated with increased odds of intrauterine adhesions (IUA) compared with two distinct control groups. This association persisted after adjustment for key confounders and remained robust in sensitivity analyses excluding patients with prior uterine surgery. Additionally, longer duration of progestin-IUD use was associated with a higher odds of IUA, suggesting a possible dose–response relationship. Interestingly, no significant association was observed between copper IUD use and intrauterine adhesions. This distinction is notable and suggests that the observed effect might relate to factors linked to the progestin-dominant hormonal milieu rather than to a foreign-body effect alone, although the low frequency of copper-IUD use in our cohort limits this comparison. Although our main analysis suggests a link between oral contraceptive use and IUA, this effect was only notable in patients with prior oral contraceptive use *and* prior progestin-IUD use, suggesting the prior progestin-IUD use is driving the association.

We observed an unexpected, but apparent protective effect of prior progestin-IUD use and subsequent endometrial polyp formation. The polyp group was originally included as a surgical comparator on the assumption that this outcome would be unrelated to the exposure. However, because our results raised the possibility that progestin exposure inversely influences membership in the polyp group, we concluded that the polyp comparator group was unable to serve as a neutral reference. As such, we designated the infertility comparator as the primary comparison and report the polyp contrast as secondary.

Our finding linking progestin-IUD exposure to IUA may be perceived as surprising, given the widespread perception of these devices as safe for uterine health and published evidence indicating a short time to pregnancy after discontinuation(3–6, 8). However, the progestin-IUD was originally approved for five years of use, and the few studies examining return to fertility after removal have largely involved durations shorter than five years in populations of both parous and nulliparous women (3–5). These studies were also relatively small (total n for all three studies <400) and combined parous and nulliparous women, so any reduction in fertility specific to nulliparous women may have been obscured. Subsequent prospective studies of conception and miscarriage rates following a range of contraceptive options have also been reassuring regarding the use of progestin-IUD’s(7, 8), although it is unclear if those with long-term use (>5 years) are adequately represented. Furthermore, studies with longer follow-up intervals that might capture complication rates subsequent to a conception or miscarriage are lacking. Thus, it is possible that our results represent a previously underrecognized association that is becoming clinically apparent as women with longer-duration use begin attempting to conceive.

The observed association between progestin-IUD use and intrauterine adhesions is biologically plausible. Progestin-IUD’s have been shown to induce significant morphological changes to the endometrium, including glandular atrophy and decidual transformation(12, 13), although these changes have been presumed to be rapidly reversible. Nevertheless, prior to full recovery, it is possible that these morphological changes may blunt the proliferative regenerative response to focal basalis injury and favor fibrotic repair over restitution. Although several mechanisms may be in play, it is known that progestins suppress endometrial matrix metalloproteinase expression, and because cyclical matrix turnover is integral to physiologic endometrial remodeling, chronic suppression could shift the balance toward matrix accumulation within an atrophic, poorly vascularized cavity (14). These pathologic pathways are supported by recent animal model studies which have linked progesterone dominance to delayed endometrial regeneration following uterine injury(15).

Future studies are needed to understand specific risk factors that may raise the risk of IUA’s following use of a progestin-IUD. Our observed dose–response pattern, wherein use of 5 years or less was associated with more modestly elevated odds while use beyond 5 years conferred markedly elevated odds, is notable. This observed duration-response association is consistent with, but does not establish, cumulative biologic effects. Longer cumulative duration could also reflect the use of more than one device in succession. The removal of a first IUD and insertion of a second into an atrophic cavity could represent a critical point of injury in a setting of limited regenerative capacity. Interestingly, we also saw a large association between longer-term progestin-IUD use in nulliparous women (more than 12-fold), whereas we did not observe any association in parous women. It is possible that parity is protective due to the robust uterine remodeling process associated with a term pregnancy, although small numbers of parous women with extended use limits the robustness of the finding.

A major strength of our study is the relatively large sample drawn from a high-volume academic fertility center, together with the use of two distinct control groups. Although groups were identified using EHR diagnostic codes, systematic chart review confirmed the accuracy of outcome, exposure, and covariate ascertainment. Several limitations should nonetheless be considered. The choice of a control group is a key determinant of effect estimates in case-control studies. Prior progestin-IUD use in our infertility comparator (11.1%) was lower than the 15.3% lifetime prevalence of hormonal IUD use among U.S. women ages 15–49 in the 2015–2019 National Survey of Family Growth (NSFG) (11). This could reflect the racial composition of our population: both our data and the NSFG show substantially lower hormonal IUD use among Asian women (8.0% nationally), who made up a substantial share of our cohort. Importantly, Asian patients comprised a similar proportion of the case and controls groups (27% and 31%, respectively), so this compositional effect would be expected to lower IUD prevalence in both groups rather than to distort the case-control contrast. We nonetheless acknowledge that any net under-representation of the exposure among controls would tend to inflate rather than attenuate the observed odds ratios. Substituting the unadjusted national benchmark (15%) for the observed control rate would attenuate the association between the progestin-IUD and IUA only modestly and nulling the association would require a control exposure prevalence approaching that of cases (≈30%). Control selection is therefore unlikely to account for associations of the magnitude observed, particularly the dose-response gradient at longer duration.

We also acknowledge that our retrospective design allows for potential misclassification and incomplete documentation, though this would tend to bias our results toward the null. Recall bias by those with IUA is a potential concern, however our contraceptive exposures were determined by intake questionnaires often completed prior to being diagnosed with IUA. Residual confounding is also possible, however reassuringly, within the infertile control group, progestin-IUD users and non-users were similar across measured characteristics apart from race/ethnicity (Supplementary Table 1), indicating that the exposure was not appreciably confounded by measured adhesion risk factors. Finally, and critically, our case–control design allows for estimates of relative associations rather than absolute or cumulative risk. Our findings therefore cannot quantify the probability that an individual progestin-IUD user will develop clinically significant intrauterine adhesions, nor the incidence of this outcome among users in the general population. This is compounded by ascertainment of cases among women presenting for fertility care and hysteroscopy, a population enriched for the outcome relative to unselected users, so the odds ratios reported here should not be interpreted as the absolute or relative risk conferred on a typical IUD user. Ultimately, further prospective studies are needed to better characterize the relationship between progestin-IUD use and IUA, particularly with respect to absolute risk. Additionally, studies should examine duration of exposure and the timing of reversibility of effects following device removal. Longitudinal studies incorporating systematic uterine cavity assessment before and after progestin-IUD use, as well as mechanistic studies examining the effects of progestins on endometrial repair and regeneration, could clarify the underlying pathophysiology.

## Data Availability

All data produced in the present study are available upon reasonable request to the authors

