## Supplementary Table 1 for "Intrauterine Adhesions and Prior use of a Progestin-releasing Intrauterine Device"

**Supplemental Table 2. Characteristics of progestin-IUD users versus non-users, within each control group**

**A. Infertile Control (n=757)**

| **Characteristic** | **Progestin-IUD users (n=84)** | **Non-users (n=673)** | **P value** |
| --- | --- | --- | --- |
| Age, years, mean (SD) | 37.1 (3.9) | 36.6 (4.1) | 0.321 |
| BMI, kg/m², mean (SD) | 24.8 (5.7) | 25.4 (5.8) | 0.550 |
| **Race/ethnicity, n (%)** |  |  |  |
| White | 50 (59.5) | 273 (40.6) | **0.001** |
| Asian | 19 (22.6) | 218 (32.4) | 0.090 |
| Hispanic | 1 (1.2) | 43 (6.4) | 0.078 |
| Black | 1 (1.2) | 15 (2.2) | 1.000 |
| Mixed | 6 (7.1) | 57 (8.5) | 0.837 |
| Unknown | 7 (8.3) | 67 (10.0) | 0.782 |
| Prior live births, mean (SD) | 0.3 (0.5) | 0.2 (0.5) | 0.472 |
| Any prior live birth, n (%) | 19 (22.6) | 130 (19.3) | 0.567 |
| Any prior spontaneous abortion, n (%) | 13 (15.5) | 102 (15.2) | 1.000 |
| Any prior induced abortion, n (%) | 7 (8.3) | 83 (12.3) | 0.374 |
| Prior D&C or MUA, n (%) | 5 (6.0) | 16 (2.4) | 0.126 |
| Prior operative hysteroscopy, n (%) | 2 (2.4) | 25 (3.7) | 0.758 |
| Prior myomectomy, n (%) | 0 (0.0) | 9 (1.3) | 0.608 |
| No prior uterine surgery, n (%) | 77 (91.7) | 624 (92.7) | 0.899 |
| Combined oral contraceptives, n (%) | 50 (59.5) | 381 (56.6) | 0.696 |

P values from independent-samples t test (continuous variables) or χ² test, with Fisher exact test for sparse cells (categorical variables). Bold P values denote P<0.05. Progestin IUD duration among users (mean [SD]): 45.8 (38.8) months. D&C, dilation and curettage; MUA, manual uterine aspiration; IUD, intrauterine device.

**B. Polyp Control(n=348)**

| **Characteristic** | **Progestin-IUD users (n=12)** | **Non-users (n=336)** | **P value** |
| --- | --- | --- | --- |
| Age, years, mean (SD) | 38.8 (5.2) | 38.4 (4.6) | 0.788 |
| BMI, kg/m², mean (SD) | 23.7 (3.2) | 24.5 (5.0) | 0.432 |
| **Race/ethnicity, n (%)** |  |  |  |
| White | 7 (58.3) | 110 (32.7) | 0.125 |
| Asian | 4 (33.3) | 151 (44.9) | 0.559 |
| Hispanic | 1 (8.3) | 24 (7.1) | 0.597 |
| Black | 0 (0.0) | 4 (1.2) | 1.000 |
| Mixed | 0 (0.0) | 3 (0.9) | 1.000 |
| Unknown | 0 (0.0) | 44 (13.1) | 0.376 |
| Prior live births, mean (SD) | 0.2 (0.4) | 0.2 (0.6) | 0.683 |
| Any prior live birth, n (%) | 2 (16.7) | 59 (17.6) | 1.000 |
| Any prior spontaneous abortion, n (%) | 2 (16.7) | 53 (15.8) | 1.000 |
| Any prior induced abortion, n (%) | 2 (16.7) | 37 (11.0) | 0.632 |
| Prior D&C or MUA, n (%) | 3 (25.0) | 32 (9.5) | 0.109 |
| Prior operative hysteroscopy, n (%) | 1 (8.3) | 34 (10.1) | 1.000 |
| Prior myomectomy, n (%) | 0 (0.0) | 8 (2.4) | 1.000 |
| No prior uterine surgery, n (%) | 8 (66.7) | 267 (79.5) | 0.285 |
| Combined oral contraceptives, n (%) | 9 (75.0) | 212 (63.1) | 0.547 |

P values from independent-samples t test or χ²/Fisher exact test, as appropriate. Bold P values denote P<0.05. Comparisons in the polyp group are limited by the small number of progestin-IUD users (n=12); P values are unstable and should be interpreted with caution. Progestin-IUD duration among users (mean [SD]): 43.0 (48.0) months.
